# Assessing Large Language Models for Oncology Data Inference from Radiology Reports

**DOI:** 10.1101/2024.05.23.24307579

**Authors:** Li-Ching Chen, Travis Zack, Arda Demirci, Madhumita Sushil, Brenda Miao, Corynn Kasap, Atul Butte, Eric A. Collisson, Julian Hong

**Affiliations:** University of California, Berkeley, Berkeley, USA; University of California, San Francisco, USA; Bakar Computational Health Sciences Institute, University of California, San Francisco, USA; Helen Diller Family Comprehensive Cancer Center, University of California, San Francisco, USA

**Author notes:** **Corresponding author information Travis Zack**, **Address:** UCSF Bakar Computational Health Sciences Institute, Box 2933, 490 Illinois St, Floor 2, San Francisco, CA 94158. Equal Contributions as first author (Li-Ching Chen, Travis Zack). Equal Contributions as last author (Eric A. Collisson, Julian Hong).

## Abstract

**Purpose:** We examined the effectiveness of proprietary and open Large Language Models (LLMs) in detecting disease presence, location, and treatment response in pancreatic cancer from radiology reports.

**Methods:** We analyzed 203 deidentified radiology reports, manually annotated for disease status, location, and indeterminate nodules needing follow-up. Utilizing GPT-4, GPT-3.5-turbo, and open models like Gemma-7B and Llama3-8B, we employed strategies such as ablation and prompt engineering to boost accuracy. Discrepancies between human and model interpretations were reviewed by a secondary oncologist.

**Results:** Among 164 pancreatic adenocarcinoma patients, GPT-4 showed the highest accuracy in inferring disease status, achieving a 75.5% correctness (F1-micro). Open models Mistral-7B and Llama3-8B performed comparably, with accuracies of 68.6% and 61.4%, respectively. Mistral-7B excelled in deriving correct inferences from “Objective Findings” directly. Most tested models demonstrated proficiency in identifying disease containing anatomical locations from a list of choices, with GPT-4 and Llama3-8B showing near parity in precision and recall for disease site identification. However, open models struggled with differentiating benign from malignant post-surgical changes, impacting their precision in identifying findings indeterminate for cancer. A secondary review occasionally favored GPT-3.5’s interpretations, indicating the variability in human judgment.

**Conclusion:** LLMs, especially GPT-4, are proficient in deriving oncological insights from radiology reports. Their performance is enhanced by effective summarization strategies, demonstrating their potential in clinical support and healthcare analytics. This study also underscores the possibility of zero-shot open model utility in environments where proprietary models are restricted. Finally, by providing a set of annotated radiology reports, this paper presents a valuable dataset for further LLM research in oncology.

## INTRODUCTION

Clinical research remains a labor-intensive task and is subject to variability and inaccuracies from human reviewers across all levels of training. The rapid evolution of artificial intelligence (AI) technologies, particularly in the realm of Large Language Models (LLMs), has ushered in a new era of possibilities in the field of clinical oncology,^1^ among them the ability to more rapidly extract clinical data from clinical text.

Most cutting-edge reports have thus far focused on the significant advancements from the proprietary GPT-3.5-turbo and GPT-4, which have demonstrated remarkable capabilities in text comprehension and generation, offering innovative approaches to interpreting complex medical data^2-4^ and the ability to perform downstream Natural Language Processing (NLP) tasks on medical documents is a budding area of current inquiry as an approximation of medical reasoning.^5,6^ This includes impressive performance in the extraction of provider notes,^7,8^ and radiology reports.^9,10,11,12^ However, these proprietary models require the transfer of information to third-party sources, which remains difficult without significant privacy and security considerations for Protected Health Information (PHI). Therefore, the comparative assessment of these proprietary models with open-source models, which can be housed and utilized locally by a single health system, is critical.

Radiology reports are text-based records produced by trained radiologists to convey information about findings within medical images.^13,14,15^ Radiographic interpretation of treatment benefits in pancreas cancer can be particularly challenging.^16,17^ Within the realm of oncology, inference of cancer trajectory from these reports is a critical task in both clinical decision-making and cohort identification in clinical research.

This study aims to bridge the gap in the current literature by evaluating the performance of both proprietary and open LLM, including generative pretrained transformer 3.5 (GPT-3.5), and GPT-4, Mistral-7B, Llama2-7B, Llama3-8B and Gemma-7B,^18-21^ in extracting and interpreting critical information from radiology reports of pancreatic adenocarcinoma patients. The focus is not only on the models’ ability to infer disease status and anatomic details but also on their capacity to extract treatment response, a crucial aspect of cancer decision making.^22,23^

## METHODS

We collected 203 radiology reports from 164 patients with pancreatic adenocarcinoma and deidentified these reports as described previously.24 Access and deidentification was performed under approval of the UCSF Institutional Review Board (IRB# 18-25163). This corpus will be made available as part of the publication for academic utilization. These included patients with new diagnoses, patients for whom surgical resection of the malignancy was completed and were presenting for surveillance imaging, and patients undergoing active treatment with systemic therapy (during which the goal of imaging is to assess treatment response). All deidentified reports, along with demographic information, are provided within a controlled access repository on physionet.org. Code is available at github.com/orpheus1234/GPT_PDAC_radiology_assessment. Figure 1 illustrates workflow. A medical oncologist experienced in treatment of pancreatic adenocarcinoma was asked to accomplish three annotation tasks for each scan: 1) Categorize disease trajectory into one of the following categories: *‘No evidence of malignancy’, ‘New Diagnosis OR disease progression’, ‘Response to treatment’, ‘Disease stability’, ‘Mixed response to treatment’, ‘Disease present but unclear trajectory of malignancy’, or ‘Unclear if cancer present on report’*, 2) Location of cancer within 15 organs/compartments and 3) Location of “Indeterminate findings/nodules that may require follow up” in these same 15 organs/compartments.

**Figure 1:**
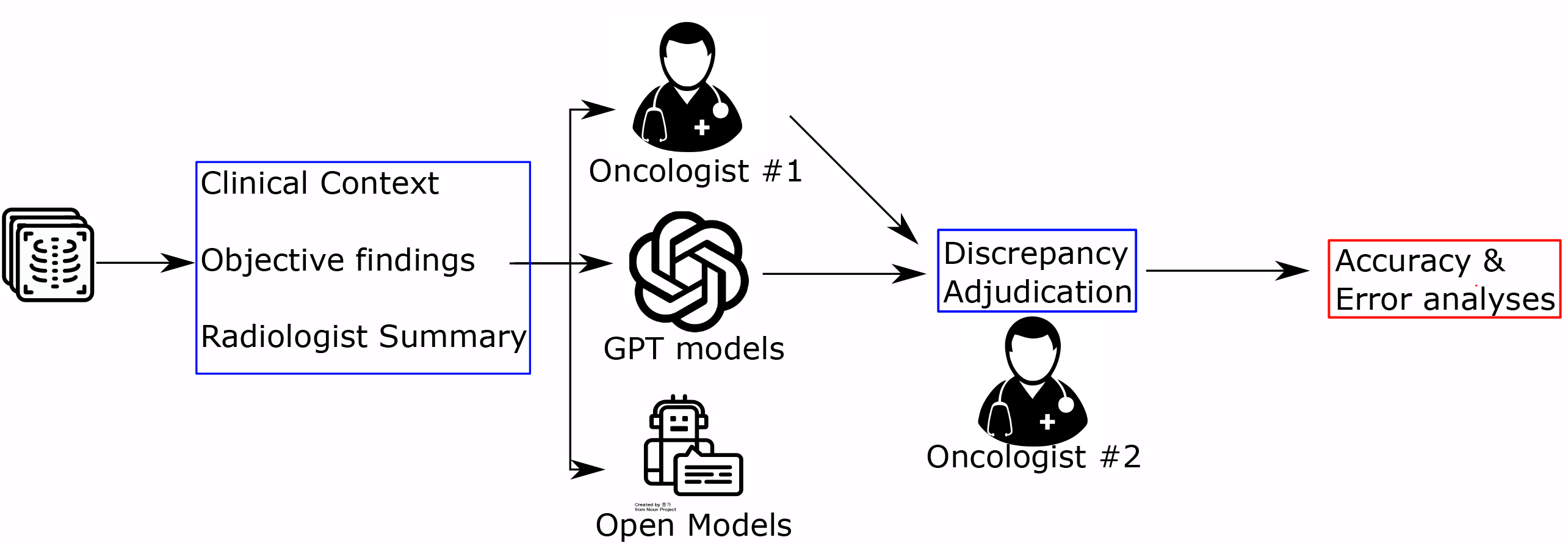
Pipeline of evaluating LLMs’ ability in oncology-related tasks with adjudication from the oncologist on discrepant results between GPT models and human annotators. The full radiology report was annotated by an oncologist for three tasks: categorizing disease trajectory, location of cancer, and location of indeterminate findings. We prompted LLMs with the reports and compared the answers with the annotations. Finally, the answers from GPT-3.5 were further adjudicated by the other oncologist not involved in the annotation process.

We then asked the LLMs to accomplish these same tasks. LLM queries were submitted via the OpenAI API via the HIPAA-compliant Microsoft Azure platform. We set the temperature parameter to the most deterministic setting, i.e., 0. We conducted separate experiments selecting the following radiology report sections.

1. The full radiology report containing A) clinical context, the date and technique of previous comparison scans (if any), B) The objective findings by organ system and C) The radiology impressions summarizing most pertinent findings
2. The sections above, excluding C) the radiologist impression
3. The sections above, excluding B) The objective findings by organ system

Prompt engineering, the construction of the language used to elicit responses from interactive LLMs, impacts model performance.^25^ Specific strategies that have shown effectiveness include asking LLM to reason through their answer^26^ and providing examples of correct output.^27^ We tested three strategies in prompt engineering: 1) Requesting that the output only contain one of the restricted pre-determined categories of answers, without any explanation of clinical reasoning. 2) Requesting it to explain the clinical reasoning of its answer, followed by one of the answer categories. Finally, we tested improvements in accuracy after providing examples of text and appropriate responses (labeled 2-shot and 3-shot). Performance was assessed by F1-micro for multiclass categorization.

To assess failure modes in GPT-3.5 performance, an oncologist not involved in the prompt design or manual annotation served as an independent adjudicator. The adjudicator assessed the reports in which there were discrepancies between human and GPT classification based on the criteria described next. Firstly, they identified whether the annotator or GPT was correct, or if either answer was reasonable in case of clinical or linguistic ambiguity. Secondly, they assessed the category of errors within the reasoning generated by the GPT model. Classes of reasoning errors could be chosen between any (or all) of following: ‘Medically inaccurate reasoning’, ‘Reasoning does not support GPT classification’ or ‘Reasoning is not supported by information in the report text’.

## RESULTS

We probed the relationship between both prompt design and report section inclusion on the performance of language models in extracting the status of a patient’s malignancy, finding a maximum accuracy of 75.5% (GPT-4, F1-micro, Figure 2, and Supplementary Figure 1). A medical oncologist determined the disease trajectory in 199 radiology reports taken from 164 patients with pancreatic adenocarcinoma (Supplementary Table 1). We evaluated the effect of prompt engineering strategies, as well as the importance of the “*Objective Findings*” vs “*Radiologist Summary*” on LLM ability to infer disease status (Figure 2). The GPT-4 model, which showed the best performance, had highest accuracy when given the full note, asked to reason through answer, and given examples of correct report/response pairs, consistent with previous reports of its significantly benefiting from being allowed to reason prior to reporting an answer.^26,28^

**Figure 2:**
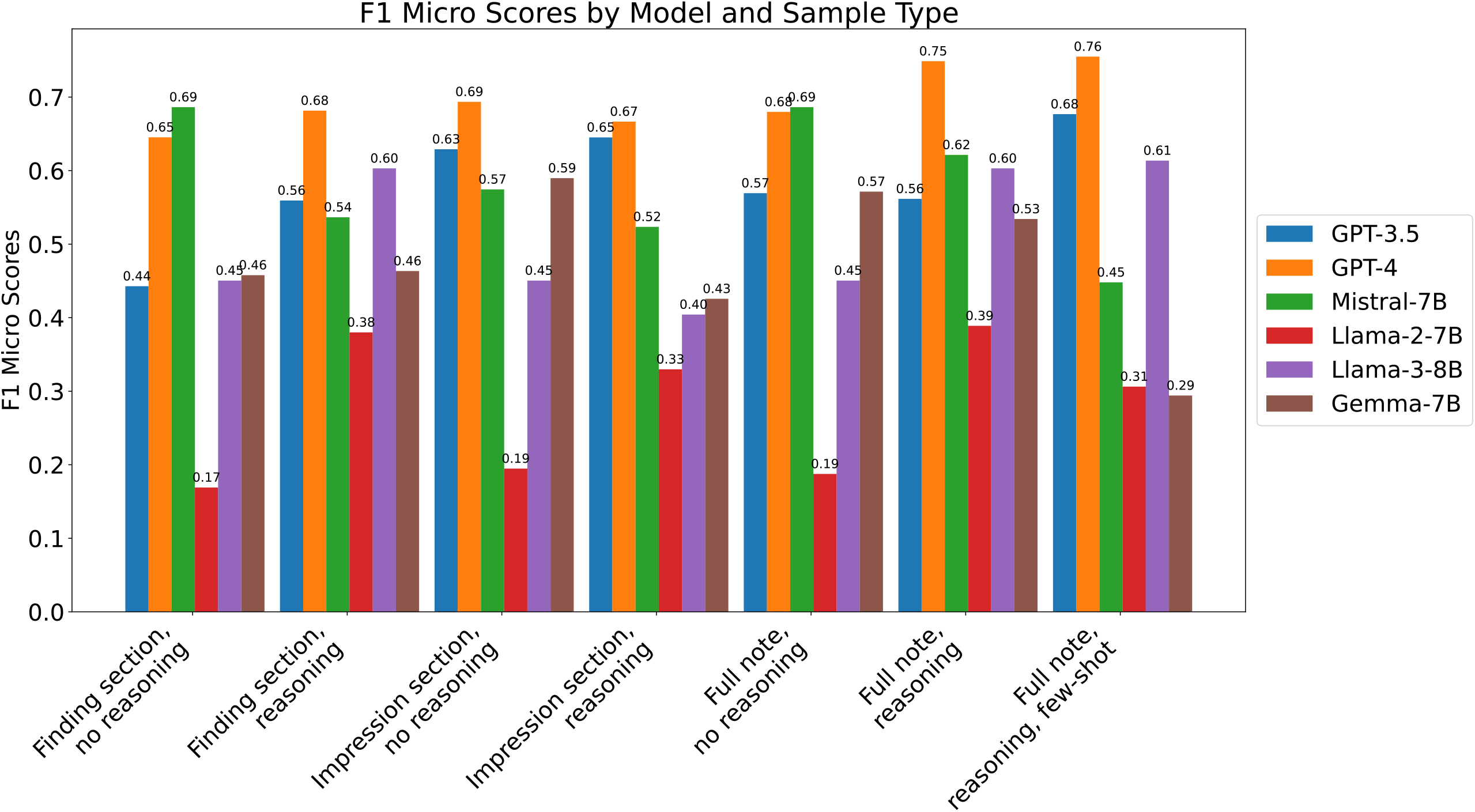
F1-micro scores of LLMs on extracting disease status from the note across different prompting strategies. F1-micro score for all models on task of identifying disease status in CT-abdomen/pelvis of patients with pancreatic cancer. We asked each model to classify each scan into one of seven states of disease (see Methods). We then tested the effect of different prompting strategies, as well as providing only specific sections of the report, on model accuracy.

Mistral-7B and Llama3-8B showed the best performance out of the open models, approaching the overall accuracy of GPT-4 (overall accuracy (Fisher exact test testing difference in accuracy between GPT-4 vs Model B) GPT-4: 75.5% vs Mistral-7B: 68.6% (0.149), GPT-3.5: 67.7% (0.09), Llama3-8B: 61.4% (0.004), Gemma-7B: 59% (0.001), Llama2-7B: 38.9% (< 0.001)) Llama3-8B shared GPT-4’s reliance on reasoning to achieve highest accuracy. However, Mistral-7B and Gemma-7B models showed their best performance when asked to give the response without reasoning, with similar performance in accuracy when given the full note versus when just provided the “*Radiologist summary*” (Figure 2). Surprisingly, when having to derive the answer directly from the “Objective Findings” section and without any intermediary reasoning step, Mistral-7B showed better performance than even GPT-4 (Figure 2, F1-micro full note, no reasoning).

A second medical oncologist adjudicated accuracy in 76 cases where disease status classification was discrepant between the original annotator and the GPT-3.5 model (Supplementary Figure 2). The expert adjudicator agreed with GPT-3.5’s inference (instead of the first annotator) in 12% of cases and felt there was sufficient ambiguity in an additional 11% of cases that both human and GPT answers were reasonable interpretations. The second oncologist was also asked to classify the root cause of the LLM misclassification by reading through the reasoning it provided (see methods). 88% of the incorrect GPT-3.5 responses were found to be due to a discordance between the reasoning (which was accurate) and the final report categorization GPT-3.5 chose, demonstrating model answers are not always concordant with reasoning generated, a frequent failure mode that has been previously reported in other contexts (Supplementary Figure 2).^29,30^

Finally, we assessed language models’ ability to parse the report by identifying the anatomic location of two clinically important entities: 1) lesions strongly consistent with cancer (“disease location”) and 2) indeterminate findings that might require further follow up (“Indeterminate findings”) (Figure 3). While GPT-4 performed significantly better than most open models at these simpler extraction tasks, Llama3-8B showed near equivalent abilities in identifying location of disease, with GPT-4 precision/recall rates (the significance level determined by Approximate randomization as described in the work of Yeh ^31^) of 0.77/0.88 vs Llama3-8B of 0.83/0.67 (0.895/< 0.0001). Overall, the open models tended to have poorer precision in the task of identifying findings indeterminate for cancer, overcalling things like post-surgical or common benign abnormalities (such as simple kidney cysts or benign liver lesions), as entities concerning for malignancy.

**Figure 3:**
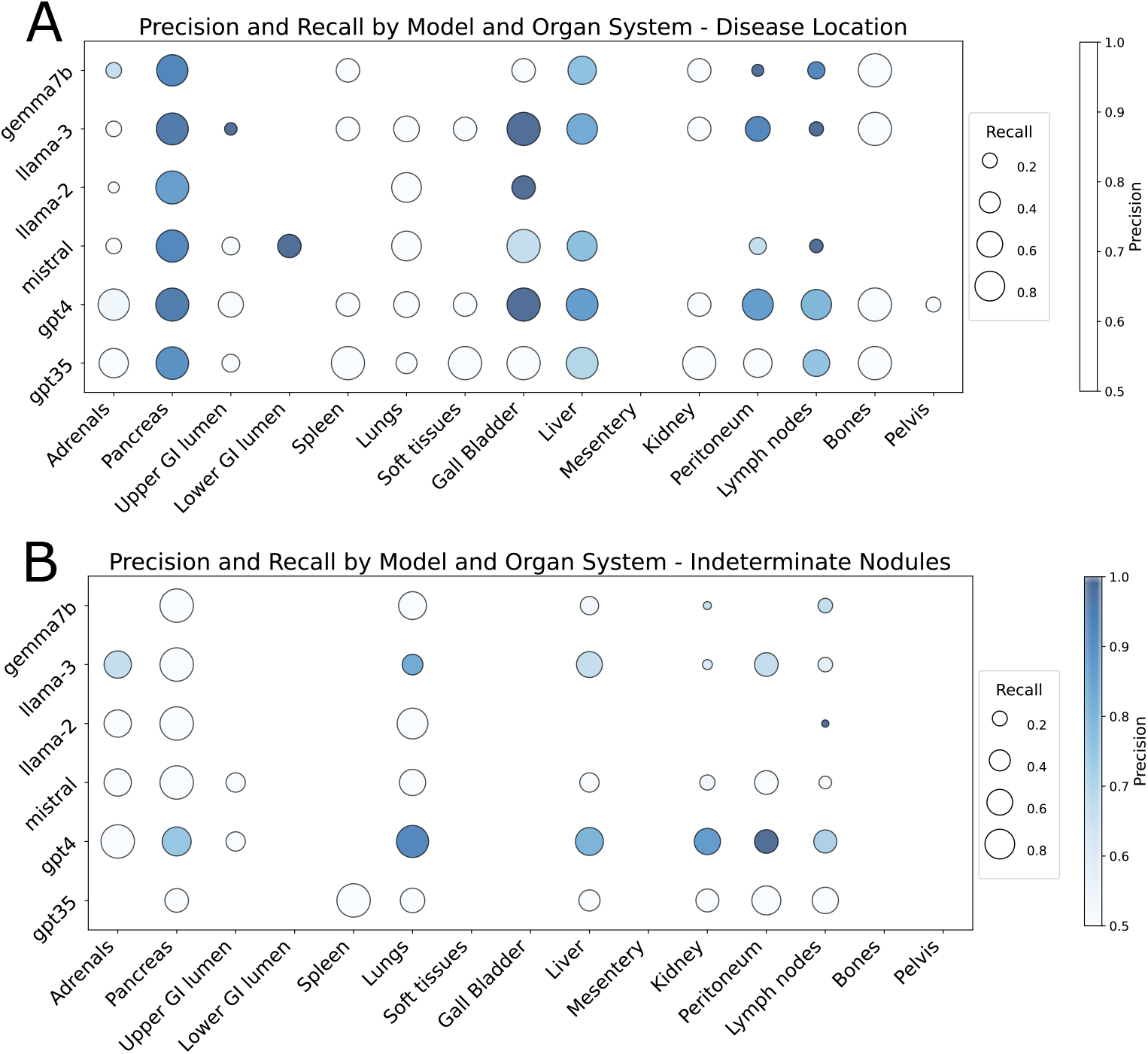
Precision and recall of LLMs in the tasks of disease location (A) and presence of findings indeterminate for cancer (B) within 15 abdominal/pelvic organs/compartments. We provided each model with the full radiology report and a list of 15 possible locations. For each report and as separate tasks, we had the model determine whether there was disease present within each of these organs (A) or whether there were indeterminate findings that could be cancer and require further follow-up (B). The size of each dot represents the model recall rate and the color represents precision.

Open models have been shown to struggle compared to OpenAI GPT models in the ability to produce an output that is properly formatted, allowing for easy answer extraction. We found that open models did require more delicate prompt engineering to ensure properly formatted output, but that with this extra effort, percentage of output that was properly formatted remained high (Supplementary Figure 3 and 4).

## CONCLUSION

Our results provide insight into the inference capacity of open-source language models in comparison to OpenAI’s GPT models in the challenging clinical task of pancreatic cancer presence and trajectory from radiology reports; a notoriously hard disease to measure and track. Our ablation and prompt engineering experiments found that the GPT models rely on the radiologist’s summarization of report when forced to classify text without generating an explanation, but this reliance can be ameliorated by allowing it to return its clinical reasoning before classification. While Llama3-8B, one of the top performing open models, similarly benefited from a human or model-produced reasoning step, Mistral-7B actually decreased in accuracy when such a step was provided/requested, suggesting chain-of-thought prompting is not a universally beneficial strategy for clinical information extraction across models. Results for GPT models were further improved by providing examples illustrating expected reasoning and answers prior to requesting text classification. Finally, we show that, while GPT performs significantly better at this task compared to most open models, open models are slowly catching up in accuracy, with the newly released Llama3-8B approaching the accuracy of GPT-4 when given the full note and allowed to reason.

We also tested these models’ ability to identify organ-specific disease involvement and presence of indeterminate findings requiring further follow up. GPT-4 shows remarkable ability to reason through its answer, exclude abnormalities that are irrelevant to the question at hand, and arrive at the correct organ involvement of disease. Llama3-8B shows comparable abilities in identification of disease location, but GPT-4 remains stronger than all other models at the more difficult task of identification of indeterminate nodules requiring further follow up. More work will need to be done under a broader set of malignancies to see whether there are organ specific features within radiology reports that may affect accuracy of these models in extracting information.

While proprietary models such as GPT-4 and MedPalm have shown incredible performance within a number of medical inference tasks^32^, they come with significant practical limitations. Firstly, they are provided as a fee-for-use model, which can be costly to run over large corpora. Sending Patient Health Information to these models is considered against HIPAA regulations unless local instances are created for each specific medical institution, which is costly and requires specific infrastructure. They also are subject to change and updates, which can make academic reproducibility challenging. Finally, they have limited ability to be trained on specific tasks, meaning their out-of-the-box performance can only be improved with prompt engineering, as opposed to fine tuning. Here we show that zero-shot open model performance can approach GPT-4 accuracy to the point of non-statistical significance in tasks of extraction an inference. However, the best choice of open model may depend on the task at hand, as in our experiments, Mistral-7B performed best overall for inference of disease status, but Llama3-8B showed stronger overall performance for location extraction tasks. These models may be further distinguished through increased performance after task-specific fine-tuning, though we note that in most settings, the training data required for this may be limited.

In summary, we have comprehensively assessed a library of currently used LLM in the ability to reason through and extract information on pancreatic malignancy from within radiologist reports and have created and provided a dataset for future academic analyses of LLM capabilities in radiology report summarization and interpretation. While it seems unlikely these tools will be used in the near future in lieu of direct study of radiology reports, the above work demonstrates the current capabilities and limitations of these models for cohort identification and information extraction for research in large real-world datasets.

## Data Availability

All data used for this study will be available on physionet at time of final peer-reviewed publication. Code, methods and prompts are contained within manuscript and supplement

## Notes

### Competing Interest Statement

The authors have declared no competing interest.

### Funding Statement

this study did not receive any funding

### Author Declarations

Access and deidentification was performed under approval of the UCSF Institutional Review Board (IRB# 18-25163).

