## Supplementary Data for "Assessing Large Language Models for Oncology Data Inference from Radiology Reports"

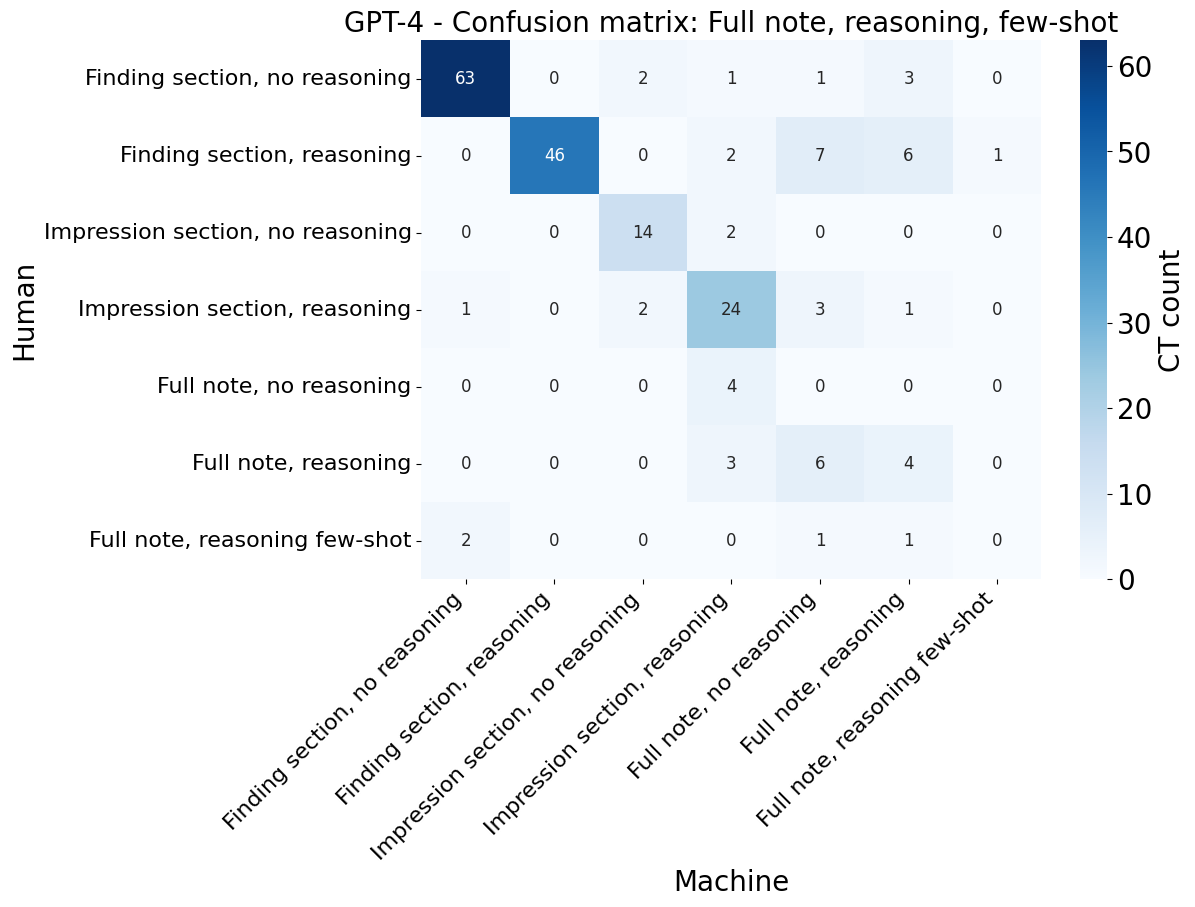

**Supplementary Figure 1: Confusion matrix for best performance text/prompt pair.** This figure illustrates the confusion matrix comparing human and GPT-4 answers when presenting GPT-4 with the entirety of the radiology report. For more confusion matrices of other models, please refer to <https://github.com/orpheus1234/GPT_PDAC_radiology_assessment/tree/main/plots/confusion_matrices>.

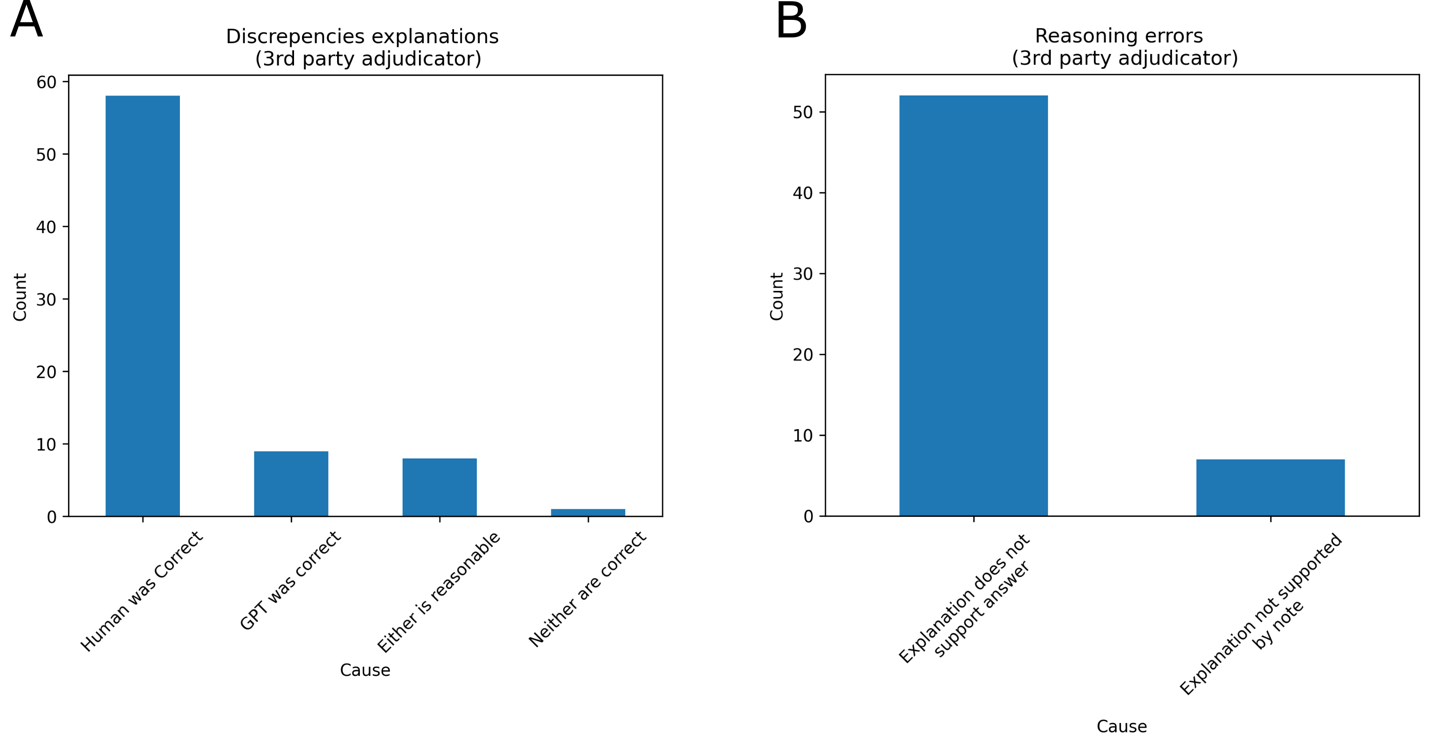

**Supplementary Figure 2. The third-party adjudicator evaluated responses from GPT-3.5 against human annotations.** A second oncologist with no role in prompt design or model development was asked to adjudicate answers provided by the first oncologist or the GPT3.5 model in a blinded fashion. Only cases where there was disagreement between human and model were considered. For each of the two responses, this annotator was asked whether answer A was correct, answer B was correct, either the answer was reasonable, or neither were correct.
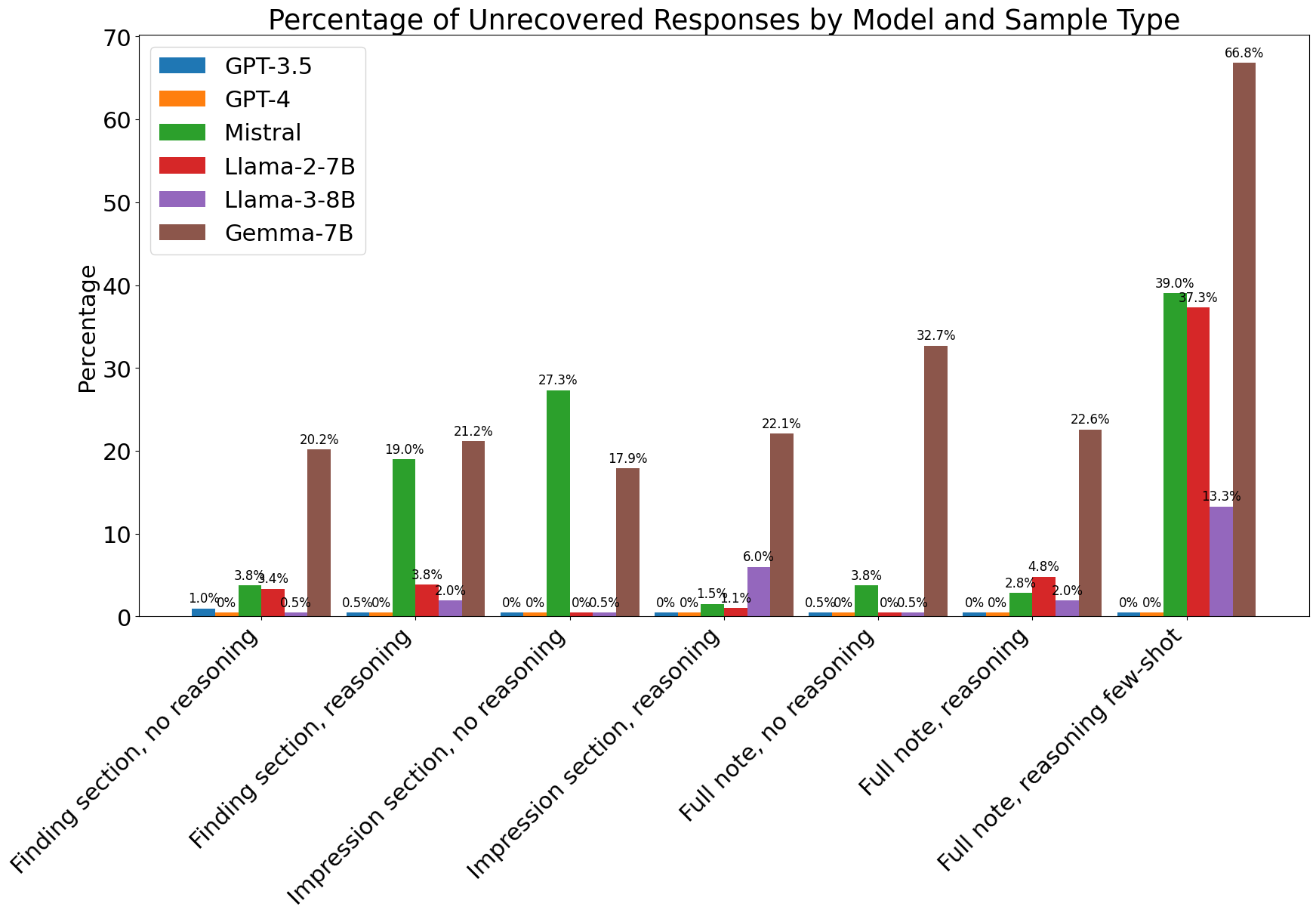

**Supplementary Figure 3. The percentage of unparsable response from the LLMs on the task of disease status.** The answer from LLM is deemed parsable by using the regular expressions, which search for entities enclosed by curly brackets as instructed in the prompt or the options listed in the prompt. GPT-4 consistently outputs the response containing a well-formatted answer, which can expedite the post-processing process in the downstream task. Compared to other open-source models, Llama3-8B has lower rates of unparsable answers across prompting strategies.

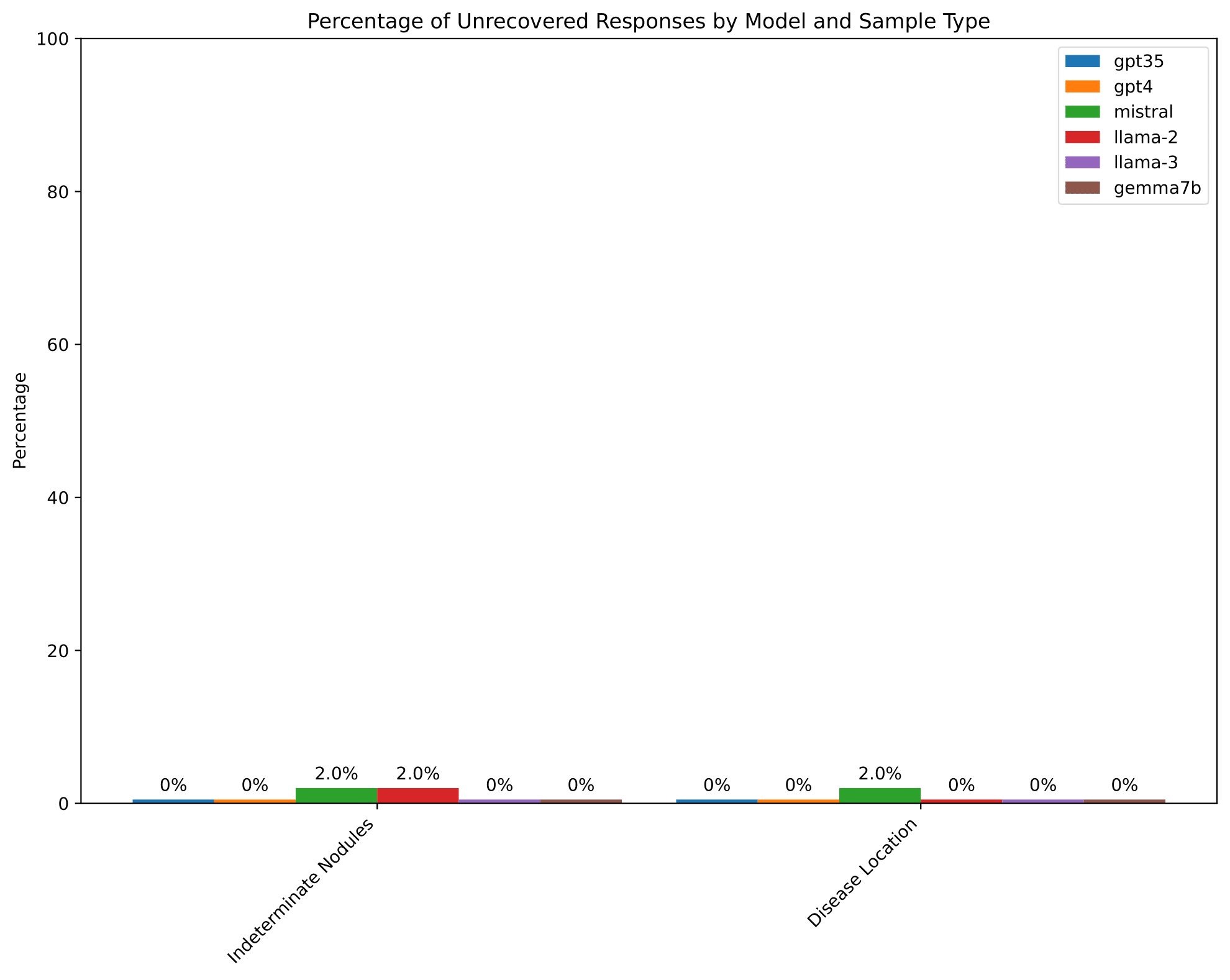

**Supplementary Figure 4. The percentage of unparsable response from the LLMs on the tasks of indeterminate findings and disease location.** All models show low rates of unparsable response in these two types of tasks.

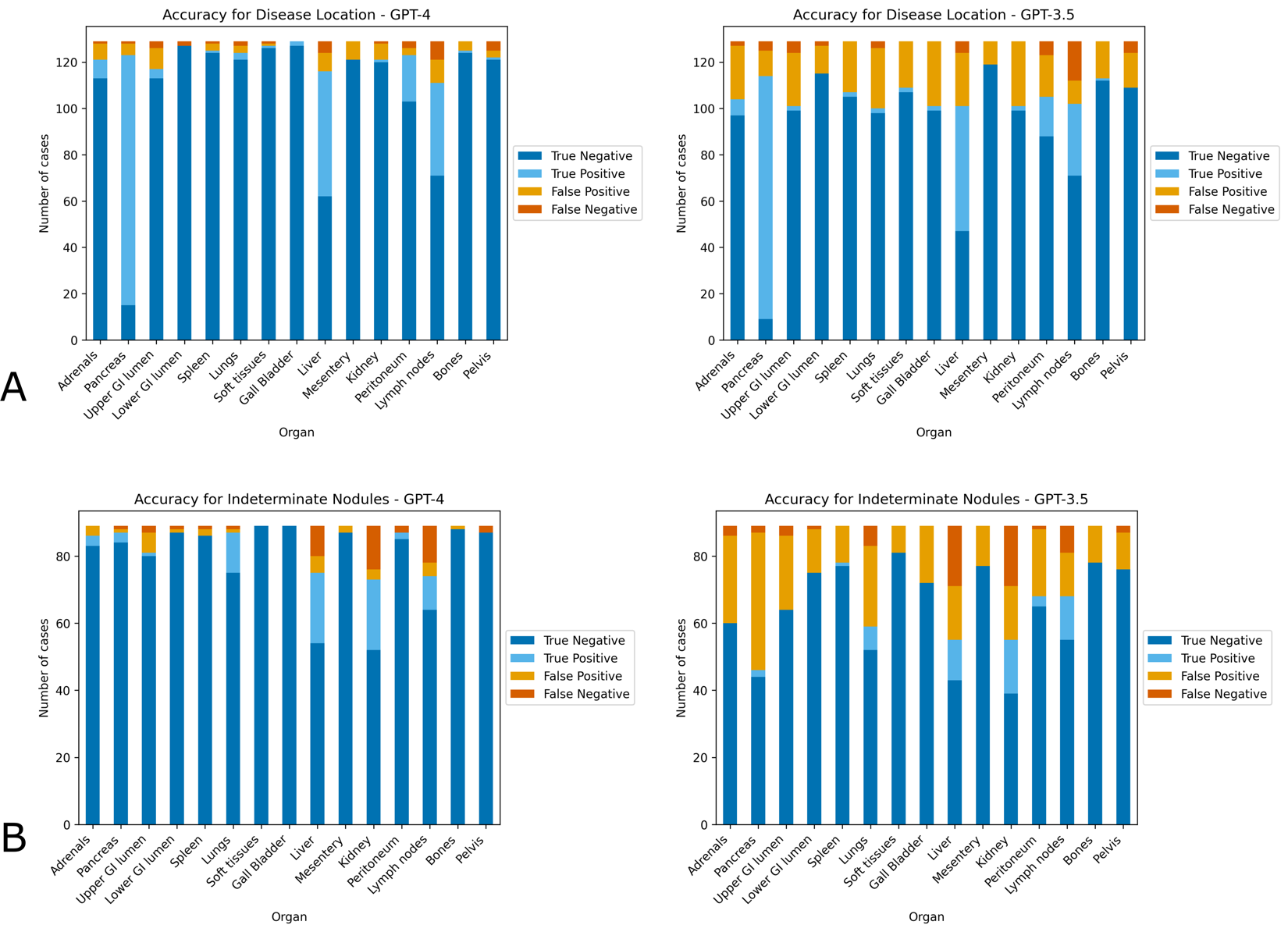

**Supplementary Figure 5: Accuracy at identification of location of disease:** GPT-4 (left) and GPT-3.5 (right) were asked to identify location of cancer (A), or presence of indeterminate findings of malignancy requiring further follow up (B). The model was asked to return results as a list of 15 pre-specified organs/compartments or report “no evidence of cancer”. In general, GPT-4 yields more accurate responses than GPT-3.5. For more results on other models, please refer to [https://github.com/orpheus1234/GPT_PDAC_radiology_assessment/tree/main/plots](https://github.com/orpheus1234/GPT_PDAC_radiology_assessment/tree/main/plots/confusion_matrices).

**Supplementary Table 1. Number of reports with annotations.** An oncologist annotated 203 radiology reports for three tasks, 1) the status of the cancer, 2) the location of the cancer, and 3) the indeterminate findings that may require further follow-up. The oncologist did not accomplish every task for every report, so this table shows the number of reports with annotations corresponding to each task.

| **Task Name** | **Number of reports with annotations** |
| --- | --- |
| Disease Status | 199 |
| Disease Location | 129 |
| Indeterminate Nodules | 89 |

**Supplementary Table 2. Number of reports for each label in each task.** The number of manual annotated classes for each task (disease status, disease location, and indeterminate nodules). For disease status, labels are chosen within seven categories mutually exclusively in the first task. Labels are chosen from 15 organs/compartments, which are not mutually exclusive, in the latter tasks. This table shows the number of reports each label has for each task.

| **Task Name** | **Label** | **Number** |
| --- | --- | --- |
| **Disease Status** | No evidence of disease | 67 |
|  | Progression | 62 |
|  | Disease Stability | 30 |
|  | Treatment response | 16 |
|  | Disease present/Unclear progression | 16 |
|  | Unclear | 4 |
|  | Mixed | 4 |
| **Indeterminate Nodules** | Adrenals | 3 |
|  | Pancreas | 4 |
|  | Upper GI tract | 3 |
|  | Lower GI tract | 1 |
|  | Spleen | 1 |
|  | Lungs | 13 |
|  | Extraperitoneal soft tissues | 0 |
|  | Gall Bladder | 0 |
|  | Liver | 30 |
|  | Mesentery | 3 |
|  | Kidney | 34 |
|  | Peritoneum | 4 |
|  | Lymph nodes | 21 |
|  | Pelvis | 2 |
|  | Bones | 0 |
|  | Other | 1 |
| **Disease Location** | Adrenals | 9 |
|  | Pancreas | 109 |
|  | Upper GI tract | 7 |
|  | Lower GI tract | 2 |
|  | Spleen | 2 |
|  | Lungs | 5 |
|  | Extraperitoneal soft tissues | 2 |
|  | Gall Bladder | 2 |
|  | Liver | 59 |
|  | Mesentery | 10 |
|  | Kidney | 2 |
|  | Peritoneum | 23 |
|  | Lymph nodes | 48 |
|  | Pelvis | 5 |
|  | Bones | 1 |
|  | Other | 1 |
|  | None | 1 |

**Task-specific prompts**

1. **Disease status with reasoning**
   a. Template to prompt large language models to elicit the disease status with reasoning based on the provided radiology note. The prompt is used for GPT-4, GPT-3.5, Mistral-7B, Llama2-7B and Gemma-7B.

Pretend you are an oncology doctor (oncologist): Below is a radiology report:

For this radiology report, we are trying to determine whether the cancer is present, and if it is present, whether it is growing, shrinking, or stable since the last time it was imaged.

Previous scan information is stored under 'COMPARISON' at the top of the report.

Note that even though the clinical history may state that this patient has pancreatic cancer, remember that the cancer could have been surgically removed, so it may no longer be present.

Think through it logically and show your work. Based on the evidence provided, is cancer present? If so, has the cancer gotten worse, improved, or remained unchanged since the last scan.

First, return the result of this reasoning. After this, please also include one of the following at the end of your report depending on your conclusion:

If there is no evidence of cancer in the scan, please return '{0: No evidence of cancer}'

If cancer was present, but there are no previous scans to compare it to, please return '{1: Cancer present, no previous scans}'

If the cancer has gotten worse or tumors have grown in size compared to prior scans, please return '{2: The cancer has progressed}'

If the cancer has shrunk or there is evidence of response to treatment, please return '{3: The cancer has responded to treatment}'

If there is cancer on the scan, but the cancer has remained mostly unchanged, please return '{4: The cancer is present and remains unchanged}'

If some parts of the cancer have grown and others have shrunk, please respond '{5: Mixed response of cancer to treatment}'

If cancer is present, but you cannot determine whether it has grown or shrunk, please respond '{6: Cancer is present but unclear if there is response to treatment}' f there is concern for cancer, but it is unclear whether cancer is present or not, please respond '{7: Unclear if cancer is present based on this report}'

Finally, if the cancer is present, but you believe this is the first time it was seen on imaging, please respond '{8: The cancer is new on this scan}'

**Note**

b. Template to prompt large language models to elicit the disease status with reasoning based on the provided radiology note. The prompt is used for Llama3-8B.

Pretend you are an oncologist. Below is a radiology report:
**Note**

For this radiology report, we are trying to determine whether cancer is present and, if so, whether it is growing, shrinking, or stable since the last time it was imaged. Previous scan information is stored under 'COMPARISON' at the top of the report. Note that even though the Clinical history may state that this patient has pancreatic cancer, remember that the cancer could have been surgically removed, so it may no longer be present. Think through it logically and show your work. Based on the evidence provided, is cancer present? If so, has the cancer gotten worse, improved, or remained unchanged since the last scan?

First, provide the reasoning. After this, please also include one of the following at the end of your report depending on your conclusion:

- If there is no evidence of cancer in the scan, please answer '{"Result": "No evidence of cancer"}'
- If cancer was present, but there are no previous scans to compare it to, please answer '{"Result": "Cancer present, no previous scans"}'
- If the cancer has gotten worse or tumors have grown in size compared to prior scans, please answer '{"Result": "The cancer has progressed"}'
- If the cancer has shrunk or there is evidence of response to treatment, please answer '{"Result": "The cancer has responded to treatment"}'
- If there is cancer on the scan, but it has remained mostly unchanged, please answer '{"Result": "The cancer is present and remains unchanged"}'
- If some parts of the cancer have grown and others have shrunk, please answer '{"Result": "Mixed response of cancer to treatment"}'
- If cancer is present, but you cannot determine whether it has grown or shrunk, please answer '{"Result": "Cancer is present but unclear if there is response to treatment"}'
- If there is concern for cancer, but it is unclear whether cancer is present or not, please answer '{"Result": "Unclear if cancer is present based on this report"}'
- Finally, if the cancer is present, but you believe this is the first time it was seen on imaging, please answer '{"Result": "The cancer is new on this scan"}'

Your Answer:

2. **Disease status without reasoning**
   a. Template to prompt large language models to elicit the disease status without reasoning based on the provided radiology note. The prompt is used for GPT-4, GPT-3.5, Mistral-7B, Llama2-7B and Gemma-7B.

Pretend you are an oncology doctor (oncologist): Below is a radiology report:

For this radiology report, we are trying to determine whether the cancer is present, and if it is present, whether it is growing, shrinking, or stable since the last time it was imaged.

Previous scan information is stored under 'COMPARISON' at top of report

If there is no evidence of cancer in the scan, please return '{0: No evidence of cancer}'

If cancer was present, but there are no previous scans to compare it to, please return '{1: Cancer present, no previous scans}'

If the cancer has gotten worse or tumors have grown in size compared to prior scans, please return '{2: The cancer has progressed}'

If the cancer has shrunk or there is evidence of response to treatment, please return '{3: The cancer has responded to treatment}'

If there is cancer on the scan, but the cancer has remained mostly unchanged, please return '{4: The cancer is present and remains unchanged}'

If some parts of the cancer have grown and others have shrunk, please respond '{5: Mixed response of cancer to treatment}'

If cancer is present, but you cannot determine whether it has grown or shrunk, please respond '{6: Cancer is present but unclear if there is response to treatment}' f there is concern for cancer, but it is unclear whether cancer is present or not, please respond '{7: Unclear if cancer is present based on this report}'

Finally, if the cancer is present, but you believe this is the first time it was seen on imaging, please respond '{8: The cancer is new on this scan}'

Please only return one of the above responses. Do NOT return any explanation of how you reached this conclusion.

**Note**

b. Template to prompt large language models to elicit the disease status without reasoning based on the provided radiology note. The prompt is used for Llama3-8B.

Pretend you are an oncologist. Below is a radiology report:
**Note**

For this radiology report, we are trying to determine whether cancer is present and, if so, whether it is growing, shrinking, or stable since the last time it was imaged. Previous scan information is stored under 'COMPARISON' at the top of the report. Note that even though the Clinical history may state that this patient has pancreatic cancer, remember that the cancer could have been surgically removed, so it may no longer be present. Think through it logically and show your work. Based on the evidence provided, is cancer present? If so, has the cancer gotten worse, improved, or remained unchanged since the last scan?

Instructions:
Please provide your answer without any reasoning or explanation. Simply include one of the following JSON-formatted responses at the end of your report, depending on your conclusion:

- If there is no evidence of cancer in the scan, please answer '{"Result": "No evidence of cancer"}'
- If cancer was present, but there are no previous scans to compare it to, please answer '{"Result": "Cancer present, no previous scans"}'
- If the cancer has gotten worse or tumors have grown in size compared to prior scans, please answer '{"Result": "The cancer has progressed"}'
- If the cancer has shrunk or there is evidence of response to treatment, please answer '{"Result": "The cancer has responded to treatment"}'
- If there is cancer on the scan, but it has remained mostly unchanged, please answer '{"Result": "The cancer is present and remains unchanged"}'
- If some parts of the cancer have grown and others have shrunk, please answer '{"Result": "Mixed response of cancer to treatment"}'
- If cancer is present, but you cannot determine whether it has grown or shrunk, please answer '{"Result": "Cancer is present but unclear if there is response to treatment"}'
- If there is concern for cancer, but it is unclear whether cancer is present or not, please answer '{"Result": "Unclear if cancer is present based on this report"}'
- Finally, if the cancer is present, but you believe this is the first time it was seen on imaging, please answer '{"Result": "The cancer is new on this scan"}'

Your Answer:

3. **Disease location**
   a. Template to prompt large language models to elicit the disease location based on the provided radiology note. The prompt is used for GPT-4 and GPT-3.5.

Below is a radiology report:
For this radiology report, we are trying to determine whether the cancer is present, and if it is present, what organs are involved.
Previous scan information is stored under 'COMPARISON' at top of report
Note that even though the Clinical history may state that this patient has pancreatic cancer, remember that the cancer could have been surgically removed, so may no longer be present.
First, please explain why you think cancer is, or is not present, and if it is present why you think certain organs are involved. Subsequently, please return any organs found to have cancer from the following list of organs in the specified format: (B) Adrenals, (C) Pancreas, (D) Upper GI tract, (E) Lower GI tract, (F) Spleen, (G) Lungs, (H) Extraperitoneal soft tissues, (I) Gall Bladder, (J) Liver, (K) Mesentery, (L) Kidney, (M) Peritoneum, (N) Lymph nodes, (O) Bones, (P) Pelvis
If there is no evidence of cancer please return: (A) No evidence of cancer
Please ONLY return organs with cancer. Do NOT report benign findings or other abnormalities.
**Note**

b. Template to prompt large language models to elicit the disease location based on the provided radiology note. The prompt is used for Mistral-7B, Llama2-7B, Llama3-8B and Gemma-7B.

Below is a radiology report:
**Note**
For this radiology report, we are trying to determine whether the cancer is present, and if it is present, what organs are involved.

Criteria for Cancer Presence:

Cancer presence within an organ can be inferred in cases where the radiologist documents that findings consistent with or suggestive of malignancy. Cancer presense should also be documented when a radiology notes a known presence of cancer in that organ that is confirmed on imaging.
Exclude findings other abnormal findings, including findings that are indeterminate and could be cancer, but could also be other benign findings according to the reading radiologist

Task Description:
First, explain your reasoning for whether there is cancer present. If so, identify the organs/compartment(s) the cancer appears to be in. Return the organs/compartments where cancer is present from the following list in the specified JSON format:
{
 "Cancer_presence": [
 {"Organ_ID": 1, "Organ_Name": "Adrenals"},
 {"Organ_ID": 2, "Organ_Name": "Pancreas"},
 {"Organ_ID": 3, "Organ_Name": "Upper GI tract"},
 {"Organ_ID": 4, "Organ_Name": "Lower GI tract"},
 {"Organ_ID": 5, "Organ_Name": "Spleen"},
 {"Organ_ID": 6, "Organ_Name": "Lungs"},
 {"Organ_ID": 7, "Organ_Name": "Extraperitoneal soft tissues"},
 {"Organ_ID": 8, "Organ_Name": "Gall Bladder"},
 {"Organ_ID": 9, "Organ_Name": "Liver"},
 {"Organ_ID": 10, "Organ_Name": "Mesentery"},
 {"Organ_ID": 11, "Organ_Name": "Kidney"},
 {"Organ_ID": 12, "Organ_Name": "Peritoneum"},
 {"Organ_ID": 13, "Organ_Name": "Lymph nodes"},
 {"Organ_ID": 14, "Organ_Name": "Bones"},
 {"Organ_ID": 15, "Organ_Name": "Pelvis"}
 ]
}

Note: Each of the above represents a SINGLE organ system. Return only the organs with cancer. Do not report benign findings or other abnormalities. Return the organs in a pipe-delimited list.

Examples:

If there is cancer in the pancreas and lungs, return: `{"Cancer_presence": [{"Organ_ID": 2, "Organ_Name": "Pancreas"}, {"Organ_ID": 6, "Organ_Name": "Lungs"}]}`
If there are no indeterminate nodules, return: `{"Cancer_presence": []}`

Your answer:

4. **Indeterminate nodules**
   a. Template to prompt large language models to elicit the indeterminate nodules based on the provided radiology note. The prompt is used for GPT-4 and GPT-3.5.

Below is a radiology report:
For this radiology report, we are trying to determine whether there are indeterminate findings that may be malignancy that require further follow up or imaging.
If there are such indeterminate nodules, we are trying to identify which organs they are present in.
We are NOT interested in findings that are obviously cancer, or relate to changes in organs where cancer is known to be present. We are also not interested in findings very likely benign.
First, explain your reasoning for why or why not there are indeterminate findings and, if so, which organ/compartment they are in. Then, using this reasoning, please return any organs/compartments that contain indeterminate findings from the following list in the specified format:'{1: Adrenals}', '{2: Pancreas}', '{3: Upper GI tract}', '{4: Lower GI tract}', '{5: Spleen}', '{6: Lungs}', '{7: Extraperitoneal soft tissues}', '{8: Gall Bladder}','{9: Liver}', '{10: Mesentery}', '{11: Kidney}', '{12: Peritoneum}', '{13: Lymph nodes}', '{14: Bones}', '{15: Pelvis}'
If there are no indeterminate nodules please return: '{0: No indeterminate nodules requiring follow up}'Please ONLY return organs with indeterminate nodules. Do NOT report benign findings or other abnormalities. Return as pipe-delimited list.

b. Template to prompt large language models to elicit the indeterminate nodules based on the provided radiology note. The prompt is used for Mistral-7B, Llama2-7B, Llama3-8B and Gemma-7B.

Below is a radiology report:
**Note**
For this radiology report, we are trying to determine whether there are indeterminate findings that may indicate malignancy and require further follow-up or imaging.

Criteria for Indeterminate Findings:

Indeterminate findings are those that do not clearly indicate malignancy but require further assessment.
Exclude findings that are obviously cancerous or related to known cancerous changes in organs.
Also exclude findings that are very likely benign.

Task Description:
First, explain your reasoning for whether there are indeterminate findings present. If so, identify the organs/compartment(s) they are in. Return the organs/compartments containing indeterminate findings from the following list in the specified JSON format:
{
 "Indeterminate_Findings": [
 {"Organ_ID": 1, "Organ_Name": "Adrenals"},
 {"Organ_ID": 2, "Organ_Name": "Pancreas"},
 {"Organ_ID": 3, "Organ_Name": "Upper GI tract"},
 {"Organ_ID": 4, "Organ_Name": "Lower GI tract"},
 {"Organ_ID": 5, "Organ_Name": "Spleen"},
 {"Organ_ID": 6, "Organ_Name": "Lungs"},
 {"Organ_ID": 7, "Organ_Name": "Extraperitoneal soft tissues"},
 {"Organ_ID": 8, "Organ_Name": "Gall Bladder"},
 {"Organ_ID": 9, "Organ_Name": "Liver"},
 {"Organ_ID": 10, "Organ_Name": "Mesentery"},
 {"Organ_ID": 11, "Organ_Name": "Kidney"},
 {"Organ_ID": 12, "Organ_Name": "Peritoneum"},
 {"Organ_ID": 13, "Organ_Name": "Lymph nodes"},
 {"Organ_ID": 14, "Organ_Name": "Bones"},
 {"Organ_ID": 15, "Organ_Name": "Pelvis"}
 ]
}

Note: Each of the above represents a SINGLE organ system. Return only the organs with indeterminate nodules. Do not report benign findings or other abnormalities. Return the organs in a pipe-delimited list.

Examples:

If there are indeterminate nodules in the pancreas and lungs, return: `{"Indeterminate_Findings": [{"Organ_ID": 2, "Organ_Name": "Pancreas"}, {"Organ_ID": 6, "Organ_Name": "Lungs"}]}`
If there are no indeterminate nodules, return: `{"Indeterminate_Findings": []}`

Your answer:
